# PAVS: A Standardized Database of Phenotype-Associated Variants from Saudi Arabian Rare Disease Patients

**DOI:** 10.64898/2026.04.05.26350189

**Authors:** Marwa Abdelhakim, Azza Althagafi, Paul N. Schofield, Robert Hoehndorf

**Affiliations:** KAUST Center of Excellence for Smart Health, King Abdullah University of Science and Technology, Thuwal, 23955, Saudi Arabia; Computer Science Department, College of Computers and Information Technology, Taif University, Taif, Saudi Arabia; Computer, Electrical and Mathematical Sciences & Engineering Division, King Abdullah University of Science and Technology, Thuwal, 23955, Saudi Arabia; Department of Physiology, Development and Neuroscience, University of Cambridge, Cambridge, CB2 3EG, United Kingdom; KAUST Center of Excellence for Generative AI, King Abdullah University of Science and Technology, Thuwal, 23955, Saudi Arabia

**Author notes:** Corresponding author: Robert Hoehndorf.

## Abstract

Genotype–phenotype databases are essential for variant interpretation and disease gene discovery. Genetic variation differs among human populations, mainly in allele frequencies and haplotype patterns shaped by ancestry and demographic history. Population-specific genotypes can influence traits and disease risk; this makes population specific characterization important. Most existing resources focus on the characterization of a population’s genetic background, but do not represent the resulting phenotypes. We have developed PAVS (Phenotype-Associated Variants in Saudi Arabia), a curated, publicly accessible database that integrates 5,132 Saudi clinical cases from four Saudi cohorts and 522 cases from analysis of a mixed-population cohort, together with 1,856 cases from the Deciphering Developmental Disorders study (DDD) and 9,588 literature phenopackets. Each case record describes patient-level phenotypes, encoded with the Human Phenotype Ontology (HPO), and links them to genomic variants, gene identifiers, zygosity, pathogenicity classifications, and disease diagnoses mapped to standardized disease terminologies. The data is represented in Phenopackets format and as a knowledge graph in RDF. Additionally, a web interface provides phenotype-based similarity search, gene and variant browsers, and an HPO hierarchy explorer. We evaluate the utility of the phenotype annotations for gene prioritization using semantic similarity. While there are clear differences to global literature-curated databases, phenotypes in PAVS can successfully rank the correct gene at high rank (ROCAUC: 0.89). PAVS addresses a gap in population-specific genotype–phenotype resources and provides a benchmark for phenotype-driven variant prioritization in under-represented populations.

## Background & Summary

Populations differ in their genetic structure as a consequence of their ancestry and demographic history. The differences in allele frequencies and haplotypes consequently affect the spectrum of Mendelian diseases they present^1, 2^. In Saudi Arabia consanguinity rates exceeding 50% increase the prevalence of autosomal-recessive disorders and create a distinct genetic architecture that global databases do not fully capture^2, 3^. Large-scale resources such as gnomAD, dbSNP, and ClinVar provide allele frequencies and pathogenicity assertions on a global scale^4–6^, but they only record coarse phenotype information (ClinVar) or aggregate phenotypes from the literature without patient-level granularity (OMIM)^7^. The Human Phenotype Ontology (HPO) annotations database (HPOA) offers structured disease–phenotype associations derived from OMIM and Orphanet records^8^, and patient-level phenotype data have been extracted from case reports through projects such as PubCaseFinder and included in the GA4GH Phenopackets repository^9, 10^. However, these resources are mainly drawn from the published literature and therefore contain phenotype profiles that are more comprehensive than those typically recorded in clinical notes^11^.

Several population-specific genotype–phenotype studies focusing on rare diseases exist, notably the Deciphering Developmental Disorders (DDD) study in the United Kingdom^12^. So far, no equivalent open resource has focused on Middle Eastern populations. The Catalogue of Transmission Genetics in Arabs (CTGA) catalogues genetic disorders in Arab populations^13^; however, CTGA uses non-standardized data formats and does not support bulk download, which limits computational reuse. Furthermore, the Saudi Human Genome Program has sequenced thousands of individuals, yet its data remain largely inaccessible for independent research. Other Middle Eastern national genome programs, including the Qatar Genome Programme^14^ and the UAE National Genome Strategy^15^, have generated large-scale sequencing datasets, but standardized genotype–phenotype data from these projects are not publicly available for computational reuse.

Phenotype data is central to variant interpretation workflows. Tools such as Exomiser^16^, Genomiser^17^, PVP^18^, and EmbedPVP^19^ rank candidate genes or variants by their phenotypic similarity to known disease profiles^20^. These methods rely on genotype–phenotype databases like the HPO database. The HPO database, however, reflects curated literature summaries rather than the incomplete, heterogeneous phenotype descriptions found in clinical practice. Consequently, the reported performance of phenotype-driven prioritization may overestimate what clinicians observe when working with real patient records.

Here, we present PAVS (Phenotype-Associated Variants in Saudi Arabia), a curated database that integrates 7,510 clinical cases (5,132 Saudi, 522 from a mixed-population cohort, and 1,856 DDD) with 9,588 literature-derived phenopackets. We standardize all records to the GA4GH Phenopackets v2.0 schema^10^ and distribute them as a SPARQL knowledge graph and a downloadable dataset. The database covers 2,389 genes and 3,528 diseases, with phenotype annotations encoded in HPO and variant coordinates in HGVS notation. Over 5,000 of the Saudi records contain genotype–phenotype associations derived from clinical notes rather than literature, providing a resource that reflects the phenotypic depth available in routine clinical practice. We evaluate the dataset through a gene prioritization experiment and show that the Saudi phenotype annotations achieve an AUC of 0.8915, well above random expectation, which confirms that even sparse clinical phenotype data carry discriminative information for gene prioritization. The lower ranking accuracy compared to literature-curated profiles is expected, as primary clinical records typically contain fewer and more general phenotype terms than published case reports.

## Methods

### Data collection

We assembled PAVS from three categories of sources: manually curated case reports from the Saudi literature, supplementary tables from large-scale Saudi cohort studies, and the reported phenotypes for genes from the DDD study cohort^12^.

We searched PubMed for publications reporting genomic variants in Saudi patients with rare Mendelian diseases, applying no date restrictions and limiting results to English-language publications. Search terms combined “Saudi Arabia”, “Saudi”, and “Arab” with “rare disease”, “Mendelian disease”, and “genomic variants”. We also performed targeted searches by screening the Google Scholar publication lists of Saudi clinical geneticists and research groups with an established focus on rare Mendelian disease in Saudi Arabia^1, 21^.

For each publication, we extracted, when available, patient-level data: demographic information (age, sex), clinical phenotypes, consanguinity status, genomic variants, zygosity, sequencing method (whole-exome, whole-genome, or gene panel), ACMG pathogenicity classification (when reported), and the PubMed identifier. Publications were included regardless of whether phenotypes were reported at the individual level or only as an OMIM disease designation, provided that variant-level information was available. Where a publication explicitly referenced cases from a prior study, we assigned the original PubMed identifier to those cases rather than the citing publication, reducing redundant entries from overlapping patient cohorts. These manually curated case reports are represented in PAVS under the prefix M and contribute 1,422 cases (Table 1).

**Table 1.**
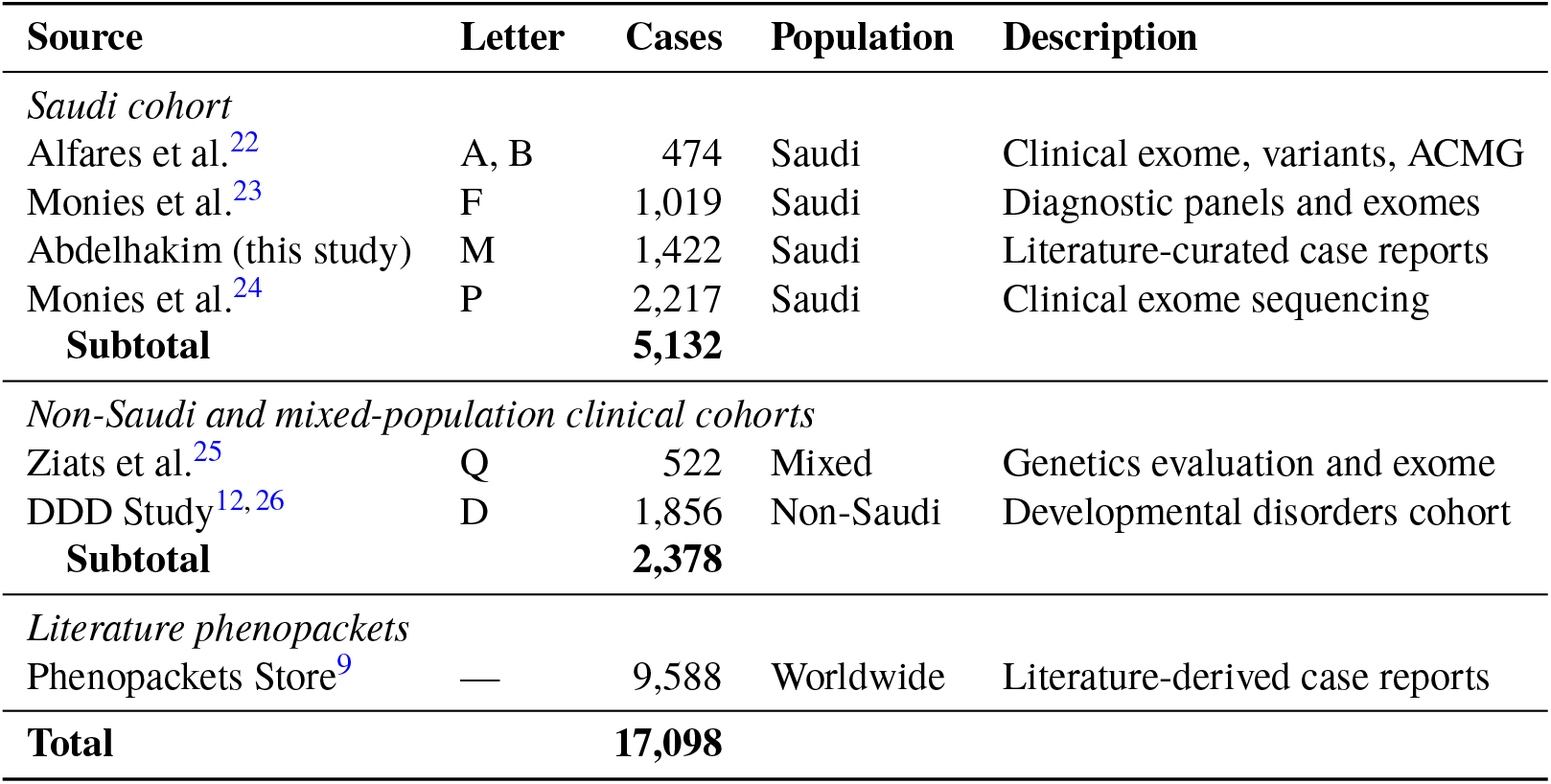
Source datasets integrated in PAVS. Letters indicate the prefix used in PAVS identifiers (e.g., PAVS:A0000001).

We integrated supplementary data tables containing phenotypes from three published Saudi cohort studies and one study from a mixed population (Table 1). For structured tables, we developed source-specific parsing scripts that handle the heterogeneous column naming and formatting conventions across publications. For each source, we designed a dedicated normalization script to extract and map the relevant fields to a common schema.

We also included 1,856 cases from the DDD study^12^, which provides gene–disease associations, semicolon-delimited HPO identifiers, and mode of inheritance. Because the DDD cohort is predominantly of European ancestry, we treat these cases as a non-Saudi comparison group.

Furthermore, we incorporated 9,588 phenopackets from the Phenopackets Store (v0.1.26)^9^, which contains case-level phenotype and variant data extracted from published case reports worldwide. These records serve as a second comparison group representing literature-derived phenotype profiles.

For the supplementary cohort tables and externally sourced datasets, including the Phenopacket Store and the DDD cohort, we did not perform systematic cross-source deduplication. Overlap between these resources and the manually curated Saudi case reports cannot be fully excluded and represents a limitation of the current dataset.

### Phenotype normalization

Clinical phenotype descriptions in the source datasets range from structured HPO identifiers to free-text clinical narratives. To account for the specific phenotype representations in the source files, we developed a custom, multi-phase phenotype matching algorithm to map these descriptions to HPO terms^8^ and simultaneously to detect negation and severity modifiers. While phenotype matching algorithms like PhenoRAG^27^ can perform this task in clinical notes and free text, the specific format of the supplementary tables required a dedicated algorithm.

The algorithm operates in two main phases. In the first phase, the complete input string is matched against the full HPO label index through three strategies applied in sequence: (1) exact string lookup against all HPO primary labels and synonyms, (2) stemmed lookup using lemmatized forms, and (3) fuzzy matching with Levenshtein distance of one. Exact matches are accepted without further validation. All non-exact matches (stemmed and fuzzy) are subject to an LLM validation step described below before being accepted.

In the second phase, the input string is segmented by punctuation delimiters and coordinated phrases are expanded (e.g., `‘‘short and malformed fingers’’` produces `‘‘short fingers’’` and `‘‘malformed fingers’’`). Each segment is processed through the same three strategies. When no match is found, the algorithm applies Aho–Corasick substring matching^28^ over the full HPO label index. As a final fallback, approximate nearest-neighbor search using SapBERT embeddings^29^ identifies semantically similar HPO terms; all results from this semantic search step are also subject to LLM validation.

For each matched span, the algorithm extracts a fixed-width word window around the match to detect negation cues and severity modifiers. Negation is detected by scanning the five words immediately preceding and following the matched span for membership in a curated negation lexicon *V*_*neg*_. This lexicon covers direct negation terms (“no”, “not”, “never”, “without”, “w/o”), clinical denial phrases (“denies”, “denied”, “ruled out”, “excluded”), absence expressions (“absence of”, “absent”, “lack of”, “free of”), result qualifiers (“negative for”, “unremarkable for”), and status-change phrases (“resolved”, “resolution of”). A phenotype is flagged as negated if any *V*_*neg*_ member occurs in either the pre- or post-span window.

Severity modifier detection scans the four words before and after the matched span for the longest matching string from the HPO modifier lexicon Λ_*mod*_, which comprises all labels and synonyms of terms in the HPO modifier sub-hierarchy (e.g., “severe”, “mild”, “progressive”, “congenital onset”). The longest match is chosen to resolve overlapping modifier phrases.

Both negation and modifier assignments are subsequently validated by the LLM. For negation, the model is asked: *“In the clinical text: ‘{context}’ — Is the phenotype ‘{term}’ expressed as absent, excluded, or not present? Reply only with: true or false*.*”* For modifiers, the model is asked: *“In the clinical text: ‘{context}’ — Is ‘{modifier}’ the severity or degree of ‘{term}’? Reply only with: true or false*.*”* An assignment is discarded if the LLM returns `false`, preventing spurious annotations from window-based false positives. The validated phenotype is then recorded with its negation status and modifier identifier in the output Phenopacket.

To guard against false-positive matches from approximate matching strategies, every candidate HPO term produced by a non-exact step is verified by a large language model (LLM) before it is added to the output. Critically, the LLM is used only as a *validator*: it is never asked to generate or suggest HPO identifiers, and it cannot introduce terms that were not already proposed by the deterministic matching stages. This design prevents hallucination errors: the LLM can only confirm or reject candidates that the algorithm has already found (same for negation and severity modifiers).

We use DeepSeek-V3^30^ (`deepseek/deepseek-chat`) via the OpenRouter API, with temperature set to 0 to ensure deterministic outputs. Two LLM operations are used.

#### Validation

For each batch of candidate terms produced by a non-exact matching step, the model receives the original clinical text and the list of candidate HPO labels and is asked to identify which labels are present or clearly implied in the text. The system prompt is: *“You are a clinical NLP expert. Determine which of the listed medical concepts are present or clearly implied in the given clinical text, including obvious typos, plurals, inflected forms, and abbreviations. Return JSON exactly as: {“valid”: [“exact label 1”, …]}”*. Only labels returned in the valid list are retained.

#### Disambiguation

When multiple HPO identifiers share the same surface label (i.e., the inverse label map returns more than one candidate), the model is asked to select the single best match given the clinical context. The system prompt is: *“You are a clinical ontology expert. Select the single best concept label that matches the clinical context. Return ONLY the exact label string, nothing else*.*”* The selected label is then resolved back to its HPO identifier.

### Variant standardization

We standardized all variant representations to HGVS notation^31^. Source-specific normalizers handle heterogeneous input formats: cDNA notation with gene context (e.g., `NM_001194998:c.2021G>T`), protein-level changes with one-letter amino acid codes (converted to three-letter HGVS p. notation), and genomic coordinates. We corrected common formatting errors including Unicode minus signs, operator spacing, and inconsistent transcript references. We also mapped zygosity annotations to the Genotype Ontology^32^ (`GENO:0000136` for homozygous, `GENO:0000135` for heterozygous, `GENO:0000134` for hemizygous, `GENO:0000402` for compound heterozygous).

For variants with sufficient coordinate information, we ran Ensembl Variant Effect Predictor (VEP)^33^ to obtain consequence predictions, SIFT^34^ and PolyPhen-2^35^ scores, gnomAD allele frequencies ^4^, and rsID identifiers. We also cross-referenced variants against ClinVar to retrieve allele identifiers and clinical significance assertions^6^. We normalized ACMG pathogenicity classifications to the five standard categories defined by the ACMG/AMP guidelines^36^ and encoded them using the OBO flatfile format^37^ as a custom ontology file (`acmg_criteria.obo`) to support ontology-based queries.

### Disease and gene mapping

We mapped disease diagnoses to OMIM^7^ and MONDO^38^ identifiers through the phenotype matching pipeline, which supports OMIM, MONDO, and Orphanet term lookup. Gene symbols were resolved to HGNC identifiers and NCBI Entrez Gene identifiers using the NCBI gene information file. For each gene, we retrieved constraint metrics (pLI, LOEUF) from gnomAD^4^, Gene Ontology^39^ functional annotations (biological process, molecular function, cellular component), tissue expression profiles from GTEx^40^ (v8), and mouse phenotype associations from MGI^41^. All annotation data are stored as literal string values in the genes named graph. The processing pipeline was implemented in Python 3.13 using rdflib 7.6.0 for RDF serialisation and pyphetools 0.9.119 for Phenopackets v2.0 assembly.

### Phenopacket generation

We converted all normalized case records to GA4GH Phenopackets v2.0 format^10^. Each phenopacket contains a subject block (identifier, sex, age), phenotypic features encoded as HPO terms with negation and modifier annotations. Disease diagnoses are recorded with OMIM or MONDO identifiers and mode of inheritance, and genomic interpretations with variant descriptors including HGVS expressions, VCF records (GRCh38), allelic state, and ACMG classification. All Saudi cases receive a `HANCESTRO:0852` (“Middle Eastern”) ancestry annotation^42^ and a `GAZ:00005279` (“Saudi Arabia”) geographic origin annotation. Source provenance is recorded in the metadata block via external references to the originating publication where available (e.g., `PMID` identifiers).

### Arabic language support

To support Arabic-speaking clinicians and patients, we generated Arabic translations of all 19,408 HPO term labels and definitions using GPT-4o^43^ via the OpenRouter API. This translation is not the official Arabic localization of HPO; it is a separate, independently curated resource distributed under CC-BY 4.0^44^ and developed mainly for the use in PAVS.

For each HPO term, the translation prompt receives the English technical name, all English synonyms, the English definition, and the names of the term’s parent classes in the HPO hierarchy (to provide disambiguation context). The model produces three fields per term: an Arabic technical name (the standard medical translation), an Arabic definition, and an Arabic layperson synonym (a simplified rendition for non-specialist readers).

The system prompt encodes domain-specific translation rules developed over multiple iterations. A mandatory glossary of 22 anatomical and morphological terms enforces consistent translations for high-frequency medical roots ***(e.g., *Atresia → ratq* 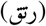, *Dysplasia → khalal tanassujī* 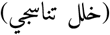, *Metaphysis → kurdũs* 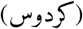. Glossary terms were selected using three complementary criteria: (1) frequency of occurrence across HPO labels, prioritizing roots appearing in a substantial proportion of terms where inconsistent translation would have the broadest ontology-wide impact; (2) error patterns identified during expert review, where any root generating inconsistent or incorrect translations constituted an empirically grounded selection candidate; and (3) systematic coverage of the major morphological and anatomical categories represented in HPO, including morphogenesis modifiers (aplasia, hypoplasia, agenesis, dysplasia), anatomical structural roots, and quantitative modifiers (hyper-, hypo-, macro-, micro-). Where available, selected Arabic technical labels were cross-referenced against established Arabic biomedical terminology resources; the WHO/EMRO Unified Medical Dictionary^45^ served as the primary source of standardized Arabic medical nomenclature, and the WHO Arabic ICD-11 browser^46^ provided secondary validation for disease-oriented terms.

Arabic sentence structure rules require stating the abnormality before the anatomical location, following the pattern [*abnormality*] + *fī* 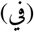 + [*location*], which reverses the typical English word order. Morphogenesis rules mandate specific prefixes for *hypoplasia* (*naqs tanassuj* 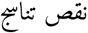), *aplasia* (*àdam at-tanassuj* 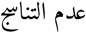), and *agenesis* (*àdam at-takawwun* 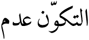), explicitly banning common alternative phrasings. A four-rule decision chain governs the translation of “Abnormal” and “Abnormality”, selecting among *shudhūdh* (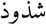, physical structural anomaly), *khalal* (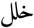 measurable functional deficit), *idtirāb* (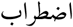 complex systemic or behavioral pattern), and *ghayr tabīì* (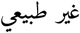, default descriptive) depending on the semantic category of the phenotype.

These rules were developed iteratively: an initial round of translations was reviewed by domain experts and Arabic speakers (A.A., M.A.), who evaluated 121 HPO terms across three quality dimensions— label accuracy, layperson synonym quality, and definition accuracy— each assessed on a binary correct/incorrect basis. Where an error was attributable solely to Arabic structural conventions rather than a translation inaccuracy, a descriptive comment was recorded in lieu of an error classification. The review identified three recurrent error types: (1) transliteration of Latin-derived terms instead of translation (e.g., “Acantholysis” rendered as a phonetic Arabic transcription rather than the standard Arabic medical term for acantholysis); (2) inconsistent morphogenesis prefixes, where terms like “aplastic” were paraphrased rather than using the canonical *àdam tanassuj* form; and (3) inconsistent rendering of the “Abnormal/Abnormality” modifier across different phenotype categories. The glossary, morphogenesis rules, and abnormality decision cascade were added to the prompt in response to these findings, and affected term categories were selectively re-translated using a substring filter mechanism (e.g., all terms containing “Abnormal” were re-translated after the four-rule cascade was introduced). Following rule implementation and selective re-translation, a second independent review assessed adherence to the updated rules and translation quality across the corrected categories.

The final translations cover 19,408 labels (100%), 19,264 definitions (99.3%; the remainder lack English definitions in the source ontology), and 19,400 layperson synonyms (100%). Translations are distributed in Babelon format for integration with the HPO internationalization infrastructure, and are also available as an OBO file for layperson synonyms.

### Datasets and ontologies

Table 2 summarizes the external ontologies and reference datasets used in PAVS, together with their versions and download locations. HPO^8^ and the HPO annotations serve as the primary source for disease–phenotype associations; MONDO^38^, GENO^32^, SIO^47^, and HANCESTRO^42^ provide supporting ontological context for disease, genotype, and ancestry annotation. gnomAD^4^, ClinVar^6^, and NCBI Gene^48^ provide variant frequency, clinical significance, and gene reference data. Gene Ontology^39^ (release 2026-01-17), GTEx^40^ (v8), and MGI^41^ provide functional annotations, tissue expression profiles, and mouse phenotype associations for gene records.

**Table 2.** External ontologies and reference datasets used in PAVS.

| Resource | Version | Download date | Source |
| --- | --- | --- | --- |
| HPO | 2026-01-08 | 2026-02-10 | https://hpo.jax.org |
| HPOA | 2026-01-08 | 2026-02-10 | https://hpo.jax.org |
| MONDO | 2026-02-03 | 2026-02-10 | https://mondo.monarchinitiative.org |
| GENO | 2020-03-08 | 2026-02-10 | https://github.com/monarch-initiative/GENO-ontology |
| SIO | 1.59 | 2026-02-10 | https://semanticscience.org |
| HANCESTRO | 2.6 | 2026-02-10 | https://github.com/EBISPOT/hancestro |
| Gene Ontology | 2026-01-17 | 2026-02-20 | https://geneontology.org |
| gnomAD | v4.1 | 2024-04-18 | https://gnomad.broadinstitute.org |
| ClinVar | 2026-02-16 | 2026-02-20 | https://www.ncbi.nlm.nih.gov/clinvar/ |
| NCBI Gene | — | 2026-02-10 | https://www.ncbi.nlm.nih.gov/gene |
| GTEx | v8 | 2026-02-20 | https://gtexportal.org |
| MGI | — | 2026-02-16 | https://www.informatics.jax.org |

### Knowledge graph generation

We transformed the phenopacket data into an RDF knowledge graph distributed across five named graphs (Table 3). The schema uses a custom PAVS ontology namespace (http://pavs.kaust.edu.sa/ontology/) with properties linking cases to phenotypes, variants, genes, and diseases. All classes and properties in the PAVS ontology are grounded in the Semantic-science Integrated Ontology (SIO)^47^. The ontology defines five classes: `pavs:Case` (subclass of sio:`SIO_000393`, patient), `pavs:GenomicVariant` (subclass of sio:`SIO_001381`, sequence variant), `pavs:GeneRecord` (subclass of `sio:SIO_010035`, gene), `pavs:DiseaseHPOAssociation` (subclass of `sio:SIO_000897`, association), and `pavs:HPOTerm` (subclass of `sio: SIO_000005`, informational entity).

**Table 3.** Named graphs in the PAVS knowledge graph.

| Graph | Triples | Contents |
| --- | --- | --- |
| cases | 175,969 | Saudi, mixed-population, and DDD case records |
| genes | 147,331 | Gene annotations (GO, GTEx tissue expression, mouse phenotypes, pLI) |
| hpoa | 1,613,546 | Disease–HPO associations |
| hpo-ic | 56,424 | HPO IC values, hierarchy, labels |
| literature | 407,246 | Literature phenopackets |

Fourteen object properties (e.g., `pavs:hasPhenotype, pavs:hasVariant, pavs:affectsGene`) are defined as sub-properties of SIO relations, and all datatype properties inherit from `sio:SIO_000300` (has value). Standard identifiers are used for all cross-references: the HPO identifiers for phenotypes, OMIM and MONDO for diseases, HGNC and NCBI Gene for genes, GENO for zygosity, and HANCESTRO for ancestry.

The knowledge graph contains 17,098 `pavs:Case` instances (7,510 PAVS cases and 9,588 literature phenopackets), 2,389 `pavs:GeneRecord` instances, and 16,692 genomic variant instances (6,145 from PAVS cases and 10,547 from literature phenopackets) across the five named graphs. The variant count for PAVS cases (6,145) is lower than the total case count (7,510) because 1,522 cases lack a variant record, either as unsolved cases or as records in which only phenotype and disease data were available without a confirmed genotype.

Disease–phenotype associations from the HPO annotations file (`phenotype.hpoa`) are loaded into the hpoa graph, providing the reference phenotype profiles used for similarity search. HPO information content values and the HPO subclass hierarchy are pre-computed and stored in the `hpo-ic` graph to support autocomplete and phenotype-driven search at query time.

Dataset metadata and versioning are managed by a dedicated `generate_metadata.py` script that produces a VoID (Vocabulary of Interlinked Datasets)^49^ description and embeds provenance and version information into the knowledge graph. The VoID file is served as `text/turtle` at the canonical well-known URL https://pavs.phenomebrowser.net/.well-known/ void via the Nginx layer, in accordance with the W3C VoID specification. It describes the five named sub-graphs, declares linksets to 28 external URL authorities, and includes a SPARQL Service Description for the public endpoint.

The dataset adopts semantic versioning^50^. The initial release is version 1.0.0. The version string is encoded in the VoID/metadata graph using three complementary properties: `pav:version` and `dct:hasVersion` record the version literal, while `owl:versionIRI` and `pav:hasCurrentVersion` dereference to a version-specific resource URI (https://pavs.phenomebrowser.net/version/1.0.0). This allows downstream consumers and FAIR validators to unambiguously identify the exact dataset snapshot they are querying.

We further defined Shape Expressions (ShEx)^51^ for each entity type to validate the generated RDF. Four shapes constrain the graph: `pavs:CaseShape` validates case records (demographics, phenotypes, diseases, variants), `pavs:GenomicVariantShape` validates variant nodes (HGVS strings, VEP predictions, population frequencies, pathogenicity scores), `pavs:GeneRecordShape` validates gene entries (constraint metrics, GO annotations, disease associations), and `pavs:DiseaseHPOAssociationShape` validates disease–phenotype associations from the HPOA.

### Semantic similarity for gene prioritization

We implemented phenotype-based gene prioritization using semantic similarity over the HPO. Given a patient’s set of HPO terms *P* and a gene’s annotated phenotype set *G* (from the HPO gene-to-phenotype file, covering 5,193 genes), we compute pairwise term similarity using either Lin’s^52^ or Resnik’s^53^ measure, then aggregate scores with the symmetric Best Match Average (BMA)^54, 55^:

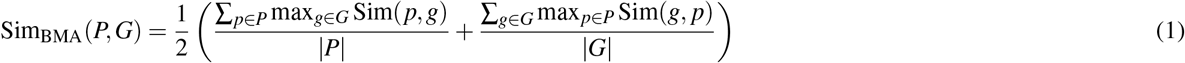

Before scoring, both the patient and gene HPO sets are expanded with all ancestor terms in the HPO hierarchy. We computed information content (IC) values using two approaches: intrinsic IC, based on HPO graph topology (IC(*t*) = − ln(desc(*t*)*/N*_total_) where *N*_total_ is the total number of terms in the HPO graph), and extrinsic IC, based on the gene annotation frequency (IC(*t*) = − ln(*f* (*t*)*/N*_genes_) where *f* (*t*) counts genes annotated to *t* or its descendants and *N*_genes_ = 5,193).

For each case in our cohort with a confirmed causative gene, we ranked all 5,193 reference genes by BMA similarity score (Equation 1) and recorded the rank of the true gene. We report Hits@*k* (the proportion of cases where the true gene ranked within the top *k*, for *k* = 1 and *k* = 10), mean reciprocal rank (MRR), and area under the ROC curve (AUC).

### Website and API

We serve the PAVS knowledge graph through a web application built with a FastAPI 0.128 backend and a React 18/TypeScript 5 frontend built with Vite 6, orchestrated via Docker Compose with an OpenLink Virtuoso triple store. At startup, the backend loads in-memory caches of IC values, HPO ancestor sets, case HPO sets, and disease labels to support sub-second similarity queries.

The web interface provides six main features: (1) phenotype similarity search, where users select HPO terms and retrieve cases ranked by Lin or Resnik BMA similarity; (2) variant lookup by gene symbol, rsID, HGVS string, or ACMG classification; (3) a gene browser listing all genes with associated Saudi case counts and detailed gene information; (4) an HPO hierarchy browser with lazy-loaded tree nodes and propagated case counts; (5) a disease browser; and (6) a SPARQL query editor for direct access to the knowledge graph. The interface supports English and Arabic through i18n localization files, including Arabic HPO term translations for all 19,408 HPO terms. All data are downloadable as individual or bundled phenopacket JSON files.

A RESTful API exposes endpoints for programmatic access, including HPO autocomplete, phenotype similarity scoring, gene and variant search, case detail retrieval, and individual or bulk phenopacket download. Full API documentation is available through an interactive OpenAPI interface.

A SPARQL endpoint at https://pavs.phenomebrowser.net/sparql provides direct programmatic access to the knowledge graph. It is powered by OpenLink Virtuoso Open Source 7^56^ and supports SPARQL 1.0 and 1.1, including federated queries. The endpoint returns results in eight formats (JSON, XML, CSV, TSV, Turtle, RDF/XML, N-Triples, and JSON-LD) and is CORS-enabled to support browser-based clients. A standard SPARQL Service Description is published at the endpoint URL.

### Data Records

PAVS contains 7,510 case records from six source datasets (Table 1), of which 5,132 are from purely Saudi cohorts, 522 are from a mixed-population cohort conducted in Saudi Arabia (Ziats et al.), and 1,856 are from the DDD cohort. Additionally, 9,588 literature phenopackets from the Phenopackets Store provide a worldwide comparison set of literature-derived case profiles.

Table 4 summarizes the database contents. The 7,510 case records span 2,389 genes and 3,528 diseases. Of these, 6,458 cases (86.0%) have at least one HPO phenotype annotation, with a mean of 7.32 HPO terms per annotated case. Regarding sex distribution, 2,192 cases are male, 2,000 are female, and 3,318 have unrecorded sex. Consanguinity is documented for 1,814 cases (24.2%). Variant zygosity is recorded for most of the 5,988 cases with a variant interpretation. Of these, homozygous variants (GENO:0000136) are the most frequent (2,690), followed by unspecified zygosity (GENO:0000137, 2,092), heterozygous (GENO:0000135, 923), and hemizygous (GENO:0000134, 102). The 2,092 cases with unspecified zygosity arise from source datasets that record variant presence without explicit zygosity information. A total of 4,391 cases (58.5%) have a solved diagnostic status.

**Table 4.** Overview of PAVS database contents.

| Category | Count |
| --- | --- |
| Total case records | 7,510 |
| Saudi cases | 5,132 |
| Mixed-population (Ziats et al.) | 522 |
| DDD cases (non-Saudi) | 1,856 |
| Literature phenopackets | 9,588 |
| Unique genes | 2,389 |
| Unique diseases (OMIM + MONDO) | 3,528 |
| Cases with $\geq 1$ HPO term | 6,458 |
| Mean HPO terms per annotated case | 7.32 |
| Cases with ACMG classification | 307 |
| Cases with VEP consequence annotation | 3,918 |
| Cases with ClinVar cross-reference | 1,224 |
| Cases with rsID | 2,667 |
| Phenopackets generated | 7,510 |
| With disease diagnosis | 2,620 |
| With variant interpretation | 5,988 |
| With phenotype features | 6,458 |
| Zygosity recorded | 5,988 |
| Homozygous (GENO:0000136) | 2,690 |
| Unspecified (GENO:0000137) | 2,092 |
| Heterozygous (GENO:0000135) | 923 |
| Hemizygous (GENO:0000134) | 102 |
| Compound het. (GENO:0000402) | 40 |
| Solved diagnostic status | 4,391 (58.5%) |

The most frequently affected genes in the Saudi cohort include ELAC2 (51 cases), ATP7B (42), TULP1 (39), ACADVL (36), and ISCA2 (33), reflecting the high burden of autosomal-recessive metabolic and neurological disorders in populations with elevated consanguinity.

All 7,510 case records are available as GA4GH Phenopackets v2.0 JSON files, a flat TSV table, and an RDF knowledge graph, individually and as a combined download via the PAVS website (https://pavs.phenomebrowser.net) and archived at Zenodo^57^ (https://doi.org/10.5281/zenodo.19311539).

The RDF knowledge graph is accessible through a public SPARQL endpoint and the PAVS web interface. The graph contains five named sub-graphs totalling approximately 2.4 million RDF triples (Table 3).

## Technical Validation

### Performance in gene prioritization

We evaluated the utility of the PAVS phenotype annotations for phenotype-driven gene prioritization by comparing the performance across three cohorts: PAVS clinical (Saudi and mixed-population, 3,088 cases with known causative genes), DDD (1,443), and literature (8,887). For each case, we ranked all 5,193 genes in the HPO gene-to-phenotype reference by BMA semantic similarity (Equation 1) and recorded the rank of the true causative gene.

Table 5 reports the results for all four IC/similarity combinations (extrinsic Resnik, extrinsic Lin, intrinsic Resnik, and intrinsic Lin); the extrinsic Resnik method consistently outperformed the others across all three cohorts. The Literature cohort achieved 57.78% Hits@1 and an MRR of 0.6542, while the DDD cohort achieved 62.02% Hits@1 and an MRR of 0.7063. In contrast, the PAVS clinical cohort reached only 3.69% Hits@1 and an MRR of 0.0676, despite maintaining a high AUC of 0.8915.

**Table 5.** Gene prioritization performance across four IC/similarity combinations using BMA semantic similarity. All metrics are computed by ranking 5,193 reference genes against each case’s HPO profile. Bold values indicate the best-performing method per cohort.

| Cohort | Method | AUC | MRR | Hits@1 (%) | Hits@10 (%) |
| --- | --- | --- | --- | --- | --- |
| Literature (n=8,887) | Extrinsic Resnik | **0.9828** | **0.6542** | **57.78** | **77.87** |
|  | Extrinsic Lin | 0.9811 | 0.6465 | 56.82 | 77.91 |
|  | Intrinsic Resnik | 0.9814 | 0.6003 | 52.18 | 73.23 |
|  | Intrinsic Lin | 0.9797 | 0.5920 | 51.60 | 72.65 |
| DDD (n=1,443) | Extrinsic Resnik | **0.9951** | **0.7063** | **62.02** | **85.45** |
|  | Extrinsic Lin | 0.9941 | 0.6841 | 60.29 | 83.99 |
|  | Intrinsic Resnik | 0.9929 | 0.6240 | 53.71 | 78.59 |
|  | Intrinsic Lin | 0.9925 | 0.6436 | 55.86 | 80.32 |
| PAVS clinical (n=3,088) | Extrinsic Resnik | **0.8915** | **0.0676** | **3.69** | **12.47** |
|  | Extrinsic Lin | 0.8870 | 0.0605 | 2.98 | 11.24 |
|  | Intrinsic Resnik | 0.8896 | 0.0585 | 2.91 | 11.24 |
|  | Intrinsic Lin | 0.8892 | 0.0558 | 2.56 | 10.56 |

The apparent contrast between these two metrics reflects a difference in what they measure. AUC quantifies the probability that the true gene ranks above a randomly chosen negative gene, averaged across all possible thresholds; it captures global ranking quality over all 5,193 reference genes. Hits@1, by contrast, requires the true gene to rank first among all candidates, which demands that the phenotype profile be specific enough to resolve the causative gene from every alternative. With a mean of only 3.8 HPO terms per case and a mean IC of 2.581, the Saudi clinical annotations carry sufficient discriminative information to place the true gene well above random expectation (AUC 0.8915), but not to resolve it as the single top-ranked candidate. A high AUC with a low Hits@1 is therefore expected when phenotype profiles are sparse: the annotations support phenotype-driven candidate shortlisting but are insufficient for reliable top-1 resolution. This outcome is consistent with the intended use of PAVS, which is to provide a population-specific genotype–phenotype resource for downstream prioritization and research, rather than a standalone gene prediction tool.

The performance gap reflects the differences in phenotypic depth. Saudi cases have a mean of 3.8 HPO terms per case (SD 2.8), compared to 19.3 (SD 15.8) for DDD and 21.5 (SD 21.9) for literature cases (Table 6). The mean IC per term is also lower in Saudi cases (2.581 vs. 3.221 for DDD and 3.482 for literature), indicating that the Saudi, clinically-derived annotations tend to use more general, higher-level HPO terms that carry less discriminative information for gene prioritization.

**Table 6.**
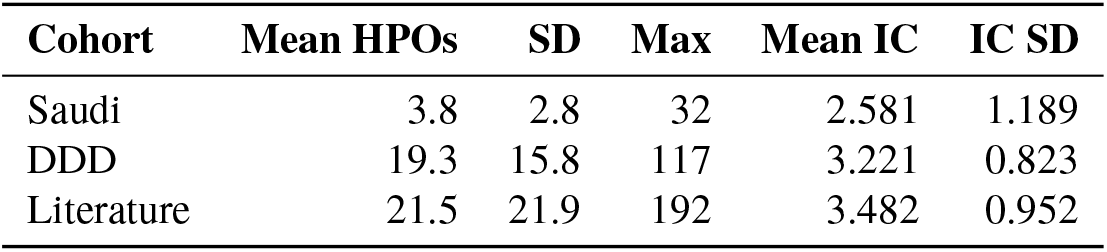
Phenotypic density and information content across cohorts.

| Cohort | Mean HPOs | SD | Max | Mean IC | IC SD |
| --- | --- | --- | --- | --- | --- |
| Saudi | 3.8 | 2.8 | 32 | 2.581 | 1.189 |
| DDD | 19.3 | 15.8 | 117 | 3.221 | 0.823 |
| Literature | 21.5 | 21.9 | 192 | 3.482 | 0.952 |

Within the Saudi cohort, performance varies substantially across source datasets. The literature-curated subset (Abdelhakim, this study), which is the only Saudi source with phenotypes curated from published case reports rather than clinical notes, achieves 8.35% Hits@1 and 21.91% Hits@10, while the remaining sources, all derived from clinical documentation, plateau between 0.66% and 2.02% Hits@1. This gap is consistent with the higher phenotypic depth that literature curation provides compared to routine clinical documentation.

We also compared four combinations of IC type (intrinsic vs. extrinsic) and similarity measure (Lin vs. Resnik); full results are reported in Table 5. Extrinsic Resnik consistently achieved the highest MRR across all three cohorts. Compared to intrinsic Resnik, extrinsic IC improved MRR by 13.2% for DDD (0.6240 → 0.7063), 9.0% for literature (0.6003 → 0.6542), and 15.6% for Saudi (0.0585 → 0.0676). The improvement from extrinsic IC is expected: because the task ranks genes, an IC measure derived from gene annotation frequencies assigns more discriminative weights to phenotype terms that distinguish between gene profiles.

### Phenopacket and RDF schema validation

We validated all generated phenopackets against the GA4GH Phenopackets v2.0 schema by checking for required fields (id, metaData), valid enumeration values for sex and progress status, well-formed HPO identifiers (matching the pattern `HP:[0-9]{7}`), and consistency between disease identifiers and OMIM/MONDO namespaces. All 7,510 phenopackets passed schema validation with zero errors. The validator produced 5,132 warnings, all caused by the `HANCESTRO:0852` (“Middle Eastern”) ancestry annotation that we include in phenotypic features for the 5,132 Saudi cases; these annotations are not part of the Phenopackets standard, but are intentionally included; they use a non-HPO identifier by design.

We further implemented a ShEx validation framework (`validate_shex.py`) that validates all RDF instances against four shape definitions: `CaseShape, GenomicVariantShape, GeneRecordShape`, and `DiseaseHPOAssociationShape`. The validator loads all generated RDF files (excluding the HPO IC graph, which contains only ontology-level triples) and evaluates each instance of the corresponding PAVS class against its shape using the PyShEx library^58^ (v0.8.1). The shape validation checks whether the expected properties and value constraints for each entity type are as specified in the PAVS schema. All validations pass.

### Data standardization quality

We assessed the quality of data standardization across four dimensions. First, we verified all assigned HPO identifiers against the HPO ontology (version 2024-08-13). The normalization pipeline always outputs valid HPO identifiers by construction: all phenotype strings are mapped through the HPO index before being written to a phenopacket, so any string that does not resolve to a current or alternate HPO identifier is rejected at normalization time. Of 47,250 HPO term assignments (present, non-excluded) across all 7,510 clinical cases, all identifiers resolve within the HPO (version 2024-08-13), yielding 100% valid HPO references.

Second, we checked HGVS variant format validity in the final stored phenopackets. Of 3,595 cDNA/RNA-level variant strings, 3,557 (98.9%) conform strictly to HGVS c. notation; the 38 exceptions use HGVS n. notation (non-coding RNA transcripts), which is valid HGVS but stored under the hgvs.c syntax tag. All 3,146 protein-level variant strings (100%) conform to HGVS p. notation, and all 3,931 genomic-level variant strings (100%) conform to HGVS g. notation with standard NC_ chromosome identifiers.

Third, all 1,838 unique disease identifiers assigned across the 7,510 clinical cases conform to standard OMIM or MONDO identifier formats (100%).

Fourth, we manually evaluated the completeness and correctness of the phenotype normalization pipeline on a random sample of 30 clinical cases drawn from the Saudi cohort. Two domain experts (M.A., P.N.S.) compared the raw clinical descriptions with the HPO terms assigned by the pipeline, grading each case as A (all phenotypes correctly captured), B (mostly correct with minor omissions), or E (substantial mapping failure). Of the 30 sampled cases, 7 lacked phenotype annotations entirely (e.g., unaffected relatives, deceased individuals with only a suspected diagnosis) and were excluded from grading. Among the remaining 23 evaluable cases, 13 (56.5%) received grade A, 9 (39.1%) grade B, and 1 (4.3%) grade E. Across these 23 cases, the pipeline mapped 61 of 69 raw phenotype mentions (88.4%) to correct HPO terms. The most common reasons for grade-B assessments were clinical findings not directly representable as HPO terms (e.g., biopsy results, disease names used as phenotype descriptions) and ambiguous abbreviations (e.g., “ASD” interpreted as atrial septal defect rather than autism spectrum disorder, “DD” not expanded to developmental delay). The single grade-E case contained only a broad diagnostic category (“Brain Malformation/Abnormal Imaging”) without specific phenotype terms, and the pipeline produced no HPO mapping. These results indicate that the normalization pipeline achieves high accuracy on well-documented clinical descriptions but, as expected, cannot recover phenotype information that is absent or described only at the level of a diagnostic category in the source data.

### Arabic translation quality

Domain experts performed an initial quality evaluation on a sample of 121 HPO terms, assessing each translation across three dimensions: technical label accuracy, layperson synonym quality, and definition fidelity.

In this initial evaluation, 77 of 121 technical labels (63.6%) were accepted without modification, 12 (9.9%) were judged acceptable but improvable, and 32 (26.4%) required revision, giving a combined acceptance rate of 73.6%. For definitions, 80 of 102 reviewed translations (78.4%) were accepted, 5 (4.9%) were acceptable but modifiable, and 17 (16.7%) required revision (combined acceptance 83.3%). The reviewers provided corrected translations for 29 terms.

The main error categories identified in this initial round were: transliteration of Latin-derived clinical terms instead of using established Arabic medical terminology (e.g., “Trichodiscoma” rendered as a phonetic transcription instead of the standard Arabic term meaning “hair disc tumor”); inconsistent use of morphogenesis prefixes, where the model paraphrased terms like “aplastic” rather than using the canonical Arabic form; and inconsistent translation of the “Abnormal/Abnormality” modifier across phenotype categories. The reviewed terms spanned multiple clinical domains, with the largest groups being skeletal/orthopedic (∼35 terms), immunological/laboratory (∼15 terms), gastrointestinal (∼10 terms), and dermatological (∼8 terms).

All issues identified in this evaluation were subsequently addressed. For systematic error categories (morphogenesis prefixes, the “Abnormal/Abnormality” modifier, transliteration of well-established medical terms), the translation prompt was updated with explicit rules mandating the correct Arabic forms, and all affected terms were re-translated by GPT-4o using the revised prompt. In rare cases where individual terms required specific corrections that did not generalize to a broader rule, the expert-provided label was adopted directly without a systematic re-translation attempt. The prompt refinements (see Methods), i.e., the mandatory glossary, morphogenesis rules, and abnormality decision cascade, are the direct result of this evaluation–correction cycle.

Reviewed translations that were accepted or corrected by the expert are marked as OFFICIAL in the distribution file (167 rows), while the remaining LLM-generated translations are marked as CANDIDATE (36,124 rows). We additionally implemented automated quality checks that flag residual Latin characters in Arabic fields, suspicious Arabic-to-English length ratios, and inconsistent prefix translations across the full 19,408-term set. These checks identified 1,227 terms (6.3%) containing Latin-script sequences, the majority of which are gene names, chemical entities, and eponyms that are conventionally retained in Latin script in Arabic clinical text; 75 terms (0.4%) with suspicious Arabic-to-English length ratios, which were manually reviewed; and multiple prefix inconsistencies (e.g. varying Arabic renderings of “Abnormal”, “Decreased”), which were standardised to ensure terminological consistency across the ontology.

### Implementation of FAIR principles

PAVS adheres to the FAIR data principles^59^. We validated compliance using the FAIR-Checker framework^60^ and describe the implementation for each principle below.

#### Findable

All case records carry globally unique, persistent identifiers following the pattern `PAVS:X0000001`, where the prefix letter encodes the source dataset. All biological entities use persistent, resolvable URIs throughout. Ontology terms use OBO PURLs (http://purl.obolibrary.org/obo/HP_ for HPO, http://purl.obolibrary.org/obo/MONDO_ for MONDO, http://purl.obolibrary.org/obo/GENO_ for GENO, etc.), which support OWL reasoning. Genes and variants use identifiers.org URIs (HGNC gene symbols, NCBI Gene IDs, dbSNP, and ClinVar). Disease entries reference OMIM directly via https://omim.org/entry/, and authors are identified by ORCID via identifiers.org. The dataset is described using a VoID (Vocabulary of Interlinked Datasets)^49^ file published at https://pavs.phenomebrowser.net/.well-known/void, providing machine-readable dataset statistics, named graph structure, and linksets to 28 external URL authorities. The SPARQL endpoint publishes a standard Service Description supporting SPARQL 1.0 and 1.1 queries with eight result formats. DCAT^61^ metadata (title, description, access URL, endpoint URL) is attached to both the dataset and the SPARQL endpoint resources. All metadata triples (207 in total) are replicated to the Virtuoso default graph to ensure discoverability by external FAIR validators that query without specifying a named graph.

#### Accessible

Data are served through three complementary channels: a public SPARQL endpoint at https://pavs.phenomebrowser.net/sparql that supports federated queries, a RESTful API with interactive OpenAPI documentation, and downloadable Phenopacket JSON files (individual and bundled). The SPARQL endpoint is powered by OpenLink Virtuoso and supports cross-origin requests (CORS), enabling federated SPARQL queries from remote endpoints and browser-based clients. The VoID file is served with appropriate content-type headers (`text/turtle`) and one-hour cache directives.

#### Interoperable

We use established standards throughout: HPO for phenotypes^8^, HGVS for variants^31^, GENO for zygosity^32^, OMIM and MONDO^7, 38^ for diseases, HANCESTRO for ancestry^42^, and GA4GH Phenopackets v2.0^10^ for data exchange. The RDF knowledge graph uses standard ontology URIs and can be queried with SPARQL 1.1^62^. All metadata is encoded using standard vocabularies: VoID^49^ for dataset statistics and linksets, DCAT^61^ for catalog metadata, Dublin Core for title, description, creator, and license, PROV-O^63^ for provenance and generation timestamps, PAV for versioning and authorship, Schema.org for search engine discoverability, FOAF for person and organization descriptions, the SPARQL Service Description vocabulary, Creative Commons for license information, and SKOS for labels. The knowledge graph links to 28 distinct external URL authorities spanning ontology repositories (OBO Foundry, W3C), biological databases (OMIM^7^, ClinVar^6^, gnomAD^4^, Ensembl^33^, UniProt^64^, Reactome^65^, DisGeNET^66^), and infrastructure registries (FAIRsharing^67^ and Wikidata^68^, where PAVS is registered as a catalogue entry).

#### Reusable

All data are released under the Creative Commons Attribution 4.0 International license (CC-BY 4.0), declared in five metadata vocabularies (`dct:license, schema:license, cc:license, dct:rights, dct:accessRights`) to ensure maximum machine readability. All source code is available under the GNU General Public License v3.0 (GPL-3.0). Ontology versions and data provenance (source file, PubMed ID) are recorded in each phenopacket. The knowledge graph includes PROV-O provenance triples recording generation activities and timestamps for each build, and PAV versioning metadata tracking version history across releases. The processing pipeline is fully documented and reproducible.

## Usage Notes

The PAVS web interface is accessible at https://pavs.phenomebrowser.net. Users can search by phenotype (selecting HPO terms for similarity scoring), by gene symbol, by variant (rsID or HGVS), or by disease name. The HPO browser allows hierarchical exploration of phenotype terms with propagated case counts. Case detail views show all associated phenotypes, variants (with VEP consequence, SIFT, PolyPhen-2, and gnomAD allele frequencies), and disease diagnoses. The interface is available in both English and Arabic, supported by a curated Arabic translation of HPO term labels and definitions.

Users can also query the RDF knowledge graph directly through the SPARQL endpoint or the built-in SPARQL editor. The endpoint supports federated queries and can be integrated with existing Semantic Web tools. Example queries are provided in the online documentation.

A REST API further provides endpoints for HPO autocomplete, phenotype similarity search, gene and variant lookup, case detail retrieval, and bulk phenopacket download. API documentation is available through an interactive OpenAPI interface.

Researchers can use the PAVS dataset to benchmark phenotype-driven variant prioritization methods on clinically derived phenotype profiles. The Saudi cohort, with its lower phenotypic depth, provides a more realistic test case than literature-derived profiles, as sparse HPO annotations better reflect the phenotypic data available in routine clinical practice.

The dataset records ancestry (`HANCESTRO`) and geographic origin (`GAZ`) for all Saudi cases, allowing population-specific analyses. The high proportion of homozygous variants among cases with a specified zygosity (69.3%; 2,690 of 3,882) reflects the consanguinity-driven genetic architecture of the Saudi population, making PAVS a valuable resource for studying recessive diseases in consanguineous cohorts.

## Code availability

All source code for data processing, normalization, phenopacket and RDF generation, the web application, and analysis scripts is archived at Zenodo^69^ (https://doi.org/10.5281/zenodo.19311276) and mirrored on GitHub (https://github.com/bio-ontology-research-group/pavs). The normalization algorithm is included in the repository. The data processing workflow, including all statistics reported in this paper, can be reproduced by following the instructions in the repository’s `PIPELINE.md` document. The primary data archive (phenopackets, RDF knowledge graph, TSV, and manually curated case data) is archived separately at Zenodo^57^ (https://doi.org/10.5281/zenodo.19311539).

## Data Availability

All 7,510 case records are available as GA4GH Phenopackets v2.0 JSON files, a flat TSV table, and an RDF knowledge graph, individually and as a combined download via the PAVS website (https://pavs.phenomebrowser.net) and archived at Zenodo (https://doi.org/10.5281/zenodo.19311539).

## Acknowledgements

This work has been supported by funding from King Abdullah University of Science and Technology (KAUST) Office of Sponsored Research (OSR) under Award No. [URF/1/5041-01-01], [REI/1/5235-01-01], [REI/1/4938-01-01], and [REI/1/5659-01-01]. This work was supported by funding from King Abdullah University of Science and Technology (KAUST) – KAUST Center of Excellence for Smart Health (KCSH) [award no. 5932], and by funding from King Abdullah University of Science and Technology (KAUST) – Center of Excellence for Generative AI [award no. 5940]. We acknowledge support from the KAUST Supercomputing Laboratory.

## Author contributions statement

M.A. curated all source datasets. A.A. conducted computational experiments and validation. P.N.S. supervised data curation and provided guidance. R.H. supervised the project, contributed code, performed validation analysis, and developed the website. All authors reviewed the manuscript.

